# What Treatment Outcomes Matter Most? A Q-study of Outcome Priority Profiles Among Youth with Lived Experience of Depression

**DOI:** 10.1101/2020.10.12.20210468

**Authors:** K. R. Krause, J. Edbrooke-Childs, H. A. Bear, A. Calderón, M. Wolpert

## Abstract

**Objective:** Over the past years, interest in youth perspectives on what constitutes an important outcome in the treatment of depression has been growing, but limited attention has been given to heterogeneity in outcome priorities and minority viewpoints. These are important to consider for person-centered outcome tracking in clinical practice, or when conducting clinical trials targeting specific populations. This study used Q-methodology to identify outcome priority profiles among youth with lived experience of service use for depression.

**Method:** A purposive sample of 28 youth (aged 16–21 years) rank-ordered 35 outcomes by importance and completed brief semi-structured interviews eliciting their sorting rationales. By-person principal component analysis was used to identify outcome priority profiles based on all Q-sort configurations. Priority profiles were described and interpreted with reference to the qualitative interview data.

**Results:** Four distinct outcome priority profiles were identified: “symptom reduction and enhanced well-being”; “improved coping and self-management”; “better understanding past and present”; and “less interference with daily life”. All four profiles prioritized outcomes related to improved mood and affect over other outcome concepts. Beyond these core outcomes, profiles differed in the level of importance assigned to learning practical coping skills, processing experiences, finding safe ways to articulate emotions, and reduced interference of depression with life and identity.

**Conclusion:** As part of a person-centered approach to care delivery, care providers should routinely engage young people in conversation and shared decision-making about the types of change they would like to prioritize and track during treatment, beyond a common core of consensus outcomes.

---

Depression is a common and serious mental health problem in adolescence, with lifetime prevalence rates of 11–16% in the United States and Europe [1–3]. Unless treated effectively, adolescent depression can have negative impacts on mental and physical wellbeing, educational attainment, employment, and income across the life course [4– 10]. There is a need to enhance the efficacy of available treatment options, as well as their effectiveness in clinical practice. Within a framework of person-centered care, such treatments should enable change in outcomes that matter to service users and their families [11, 12], and these outcomes should be tracked in clinical trials and routine practice, to ensure that treatment efficacy and effectiveness are appraised in relation to service user priorities. Yet, outcome priorities of young people experiencing depression have received limited attention thus far. Measurement approaches have historically been determined with minimal youth input, and focused on symptomatic change [13–15]. Consultations with young people in mental health contexts suggest they value additional outcomes such as improved coping skills, affirming their identity, increased autonomy, feeling more connected with others, improved family relationships, or better school functioning [16–21]. Youth have expressed a preference for outcome measurement that is meaningful and tailored to their individual priorities [22–24].

Over the past years, initiatives have emerged that aim to form consensus between researchers, clinicians, youth, and families on the outcomes considered most important. The International Consortium for Health Outcome Measurement (ICHOM) convened a multi-stakeholder working group to devise a standard set of outcomes to be measured for all children and young people with depression or anxiety treated in routine care settings worldwide [25]. The standard set recommends tracking outcomes in the domains of symptom change, functioning, and suicidal thoughts and behavior, as a minimum. A second initiative is led by the International Network for Research Outcomes in Adolescent Depression Studies (IN-ROADS) and aims to identify core outcomes for youth depression trials [26].

These initiatives engage young people in the identification and selection of outcomes and push the field towards accommodating different stakeholder perspectives on what matters most. Yet, the focus is typically on identifying a minimum set of outcomes that all stakeholder groups can agree upon. Consensus-building methodology is geared towards creating common ground, rather than investigating differences in perspective. Similarly, existing qualitative research into notions of “good outcomes” has focused on identifying dominant themes, rather than exploring heterogeneity or minority viewpoints [16, 20, 21, 27]. Within a framework of person-centered care, the latter are important to consider if outcome measurement is to be personalized and tailored to individual needs, beyond a common core set of consensus outcomes.

A method that is tailored to the systematic study of distinctive viewpoints is Q-methodology [28, 29]. As part of a Q-methodological study, researchers assemble a set of items that represent the current discourse on a phenomenon of interest. Participants are then invited to sort these items according to a pre-defined ranking scheme [30]. Patterns of similarity and divergence in the resulting *Q-sorts* are analyzed using inverted or “by-person” factor analysis, where factors are based on the correlations between the overall Q-sort configurations produced by participants, rather than the correlation between individual Q-sort items [31]. The viewpoints identified are described and interpreted qualitatively. Q-methodology combines the systematic study of subjectivity with the transparency and rigor of quantitative analysis [32, 33]. The technique moves beyond eliciting a majority view by aiming to identify a range of distinctive viewpoints, with special attention to minority experiences [34, 35]. Q-methodology has been previously deployed with adolescents in health contexts, examining preferences for hospital care for chronic physical conditions, antidepressant side effects [36], reasons for treatment non-adherence in renal and kidney transplant patients [37, 38], and attitudes towards health-related lifestyles amongst non-clinical youth [39]. To the best of our knowledge, no Q-study has yet examined outcome priorities in relation to adolescent depression.

This study aims to apply Q-methodology to the study of outcome priority profiles amongst young people with lived experience of mental health service use for depression. This study was part of a wider Q-methodological research project into outcome priorities for adolescent depression, which included parallel investigations with mental health practitioners in the United Kingdom (UK) and in Chile.

## Method

### Participants

Q-methodological studies aim to identify and describe viewpoints in depth, rather than assert their prevalence in a reference population [40]. As examinations of subtle patterns of meaning become difficult with increasing data volumes [33, 41], Q-studies typically include between 20 and 50 purposively sampled participants. In inverting sample size guidelines for traditional factor analysis, it is recommended that the number of participants should not exceed the number of items to sort [42, 43]. As the present Q-study employed a 35-item Q-set (see below), we aimed for a maximum participant sample of 30 young people, but used a principle of saturation: once a minimum of 25 participants had been recruited, additional interviews were conducted until no substantially new viewpoints were articulated [44].

Given limited evidence on the demographic and clinical characteristics influencing youth outcome priorities, sampling was not hypothesis-led. Instead, we aimed to represent varying depression histories and socio-economic profiles. Adolescents were eligible if aged 12 to 21 years, with lived experience of accessing mental health support for depression (as per self-report). Co-occurring presenting problems (including neurodevelopmental disorders) were not an exclusion criterion, if young people were able to complete the research tasks in a self-directed manner. However, youth known to be experiencing acute suicidal ideation or psychosis at the time of the research were excluded for safeguarding reasons. Using convenience sampling [45], we advertised recruitment calls through the networks of youth mental health charities in England; the University College London Psychology Subject Pool; social media; and by soliciting youth peer support groups.

The Q-sort was completed by a volunteer sample of 28 adolescents. As intended, the sample was diverse with regards to socio-demographic profiles, experiences of service use, and self-reported co-occurring mental health difficulties (see Table 1). Eighteen youth (64%) identified as female, nine identified as male (32%) and one identified as non-binary. The mean age was 18.7 years. On average, each young person reported to have experienced four co-occurring presenting problems in addition to depression, with anxiety being the most common (82%), followed by sleep problems (64%), self-harm (61%) and eating disorder (57%). Around half were receiving ongoing treatment (43%) while the remainder had received treatment in the past. Half of the sample (54%) had received more than one cycle of treatment. One in four youth had been admitted to emergency care in relation with their depression, and three (11%) had accessed inpatient care. The majority (86%) had received individual therapy or counseling, and more than half had been prescribed medication as part of their prior or ongoing treatment (57%).

**Table 1.** Demographic Characteristics.

| Variable | Overall |
| --- | --- |
| *N* | 28 |
| Gender |  |
| Female | 18 (64%) |
| Male | 9 (32%) |
| Non-binary | 1 (4%) |
| Mean age (SD) | 18.7 (1.8) |
| Treatment history (based on self-report) |  |
| Treatment ongoing (vs. ended) | 12 (43%) |
| Repeated cycles of treatment (vs single cycle) | 15 (54%) |
| History of admission to emergency care | 7 (25%) |
| History of inpatient care | 3 (11%) |
| Average number of co-occurring difficulties (SD) | 4.1 (2.4) |
| History of co-occurring presenting problems (based on self-report) |  |
| Anxiety or phobia | 23 (82%) |
| Sleeping problems | 18 (64%) |
| Self-harm | 17 (61%) |
| Eating problems | 16 (57%) |
| Neurodevelopmental disorder | 10 (36%) |
| Anger and violent behavior | 6 (21%) |
| Obsessions or compulsions | 5 (18%) |
| Substance use | 5 (18%) |
| Psychosis | 5 (18%) |
| Trauma | 4 (14%) |
| Types of treatment received (based on self-report) |  |
| Individual psychotherapy or counseling | 24 (86%) |
| Medication | 16 (57%) |
| Family therapy | 13 (46%) |
| Group therapy | 8 (29%) |

### Procedure

We compiled a candidate pool of outcomes through two interdisciplinary workshops involving young people; an interdisciplinary group of researchers (including psychologists, psychometricians, social scientists, and philosophers); a systematic literature review of treatment outcome studies for adolescent depression [15]; and qualitative content analysis of 102 semi-structured interviews with adolescents, their parents, and therapists following three types of psychotherapy for depression [16]. This process identified 73 possible outcomes, across the eight domains of *symptoms, self-management, functioning, personal growth, relationships, feeling seen and seeing differently, youth well-being*, and *parental support and well-being*. Outcome descriptions were refined and harmonized, and redundant items removed or collapsed [42]. Using Fisherian balanced block design, we selected four to five outcomes from each domain [46, 47] to obtain a 35-item Q-set. While it is recommended that Q-sets include between 40 and 80 items, smaller sets have been favored for use with children and young people [35, 37, 39, 48]. An early draft of the Q-set was piloted with two youth advisors, and a close-to-final version was reviewed for the clarity of the language and concepts by an adolescent volunteer. After final adjustments, each outcome description was printed on a separate numbered card for sorting.

We invited participants to rank-order the 35 outcome descriptions according to a quasi-normal distribution, using a sorting grid with a 9-point scale of importance (from +4 *most important*, to −4 *least important*, see Figure 1). Following completion, participants answered open-ended questions about the rationale for their outcome prioritization, and whether any important outcomes were missing [49]. These brief post-sort interviews were recorded and transcribed verbatim. Participants also completed a brief demographic questionnaire capturing their self-reported history of depression and co-occurring mental health problems. Half of the participant sample (*n* = 14) completed the card sorting at individual appointments; the remainder completed the task individually at a peer support group meeting. All post-sort interviews were conducted in a confidential one-on-one setting. Adolescents were remunerated for their time with £10 gift vouchers, and reimbursed for travel expenses.

**Figure 1.**
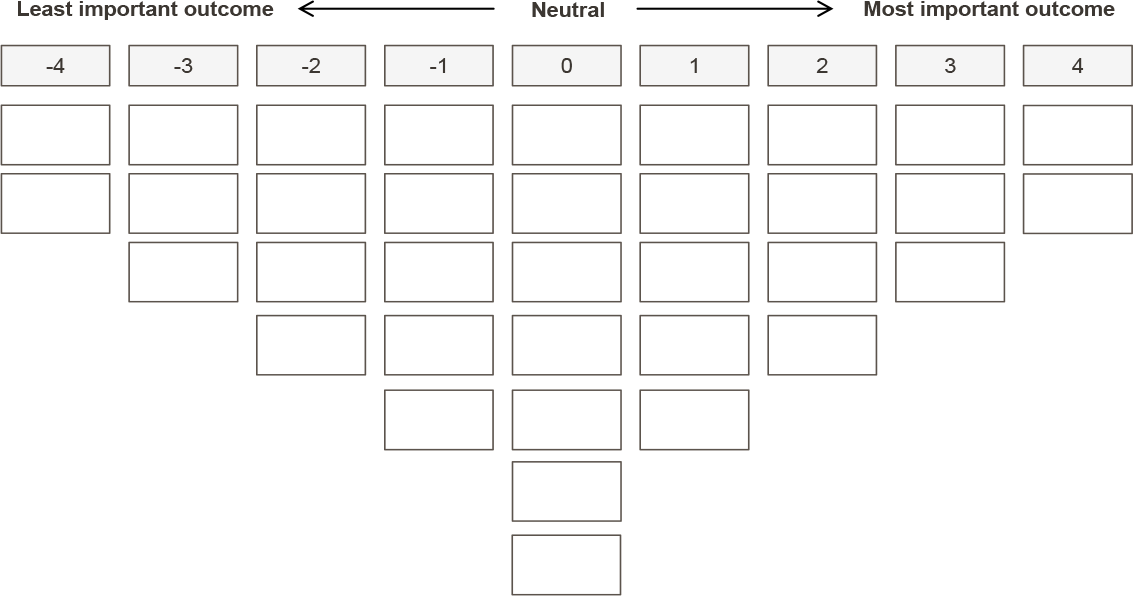
Sorting Grid Used by Participants. *Note*. The sorting grid provides 35 empty spaces—one for each item card.

### Statistical Analysis

Clusters of adolescents whose Q-sorts were highly correlated were identified via inverted (or “by-person”) principal component analysis (PCA), using the dedicated software PQMethod [50]. We identified the most suitable component solution by considering the scree plot of Eigenvalues, the shared variance explained, the number of Q-sorts loading significantly on only one principal component, and the correlation between component scores. To improve model fit, the unrotated correlation matrix was first subjected to Varimax rotation [51] to identify a solution that would mathematically maximize the variance explained [42], and small adjustments via hand rotation were then applied to increase the number of participant Q-sorts loading significantly on exactly one of the components [42].

For each extracted principal component (i.e., outcome priority profile), an ideal-typical Q-sort was generated by averaging the outcome rankings across all participant Q-sorts associated with this component, using the correlation coefficients as weights [39]. The ideal-typical Q-sort illustrates how an archetypical profile representative would have rank-ordered the 35 outcomes [42]. Outcome profiles were interpreted with reference to this ideal-typical Q-sort, and by drawing on the qualitative data collected through post-sort interviews, which were analyzed using qualitative content analysis [52].

## Ethical Considerations

This study was approved by the ethics review committee of University College London in March 2018 (UCL REC REF: 10567/002), and performed in accordance with the ethical standards of the committee and with the 1964 Helsinki declaration and its later amendments or comparable ethical standards. All participants were above the age of 16 and provided informed written consent. All interview data were anonymized, and names replaced with pseudonyms.

## Results

By-person PCA identified four outcome priority profiles that explained 48.7% of the common variance in youth Q-sorts. Inter-component correlations ranged from 0.05 to 0.33, without reaching statistical significance at *p* < 0.01, suggesting that distinct preference profiles had been extracted. Two Q-sorts did not load significantly on any principal component; and one was confounded (i.e., had significant loadings on two components). These Q-sorts were not considered for further analysis [42]. The rotated component matrix and loadings are shown in Table 2. Hereafter, we describe each of the four outcome priority profiles. We refer to the ideal-typical Q-sorts (see Table 3) by providing statement numbers and ranks in parentheses (e.g., item 3, rank +3), and draw on the post-sort interviews for interpretation.

**Table 2.** Component Loadings for Each Outcome Priority Profile.

| Outcome Priority Profile | Participant Q-sort <sup>a</sup> | Component Loadings for Each Profile |  |  |  |
| --- | --- | --- | --- | --- | --- |
|  |  | A | B | C | D |
| Profile A: Symptom reduction and enhanced well-being | Becca (17 yrs) | **.80** | .35 | .14 | .13 |
|  | Dylan (19 yrs) | **.79** | .18 | -.09 | .23 |
|  | Ellie (17 yrs) | **.69** | .09 | .18 | -.19 |
|  | Soraya (21 yrs) | **.62** | .16 | -.04 | -.11 |
|  | Samuel (17 yrs) | **.61** | .30 | .37 | .25 |
|  | Josh (18 yrs) | **.49** | .44 | .33 | .07 |
| Profile B: Improved coping and self-management | Melody (20 yrs) | -.28 | **.77** | .14 | -.04 |
|  | Adam (17 yrs) | .09 | **.68** | -.39 | .24 |
|  | Ameera (16 yrs) | -.05 | **.63** | .30 | .13 |
|  | Jacob (21 yrs) | .37 | **.54** | .10 | -.43 |
|  | Hannah (17 yrs) | .26 | **.52** | -.24 | -.02 |
|  | Liam (21 yrs) | -.07 | **.51** | .16 | .00 |
|  | Taylor (21 yrs) | .08 | **.47** | .28 | .36 |
|  | Boris (21 yrs) | .26 | **.44** | .32 | .16 |
| Profile C: Better understanding past & present | Lauren (20 yrs) | -.01 | .06 | **.74** | .05 |
|  | Chelsea (17 yrs) | .04 | -.12 | **.67** | .35 |
|  | Imogen (21 yrs) | .03 | -.02 | **.61** | -.20 |
|  | Liz (18 yrs) | -.23 | .15 | **.53** | -.04 |
|  | Connor (21 yrs) | -.03 | .44 | **.51** | .02 |
|  | Jade (19 yrs) | .39 | .28 | **.50** | -.22 |
|  | Chloe (16 yrs) | .26 | .33 | **.49** | -.18 |
|  | Amber (18 yrs) | .18 | .29 | **.44** | .32 |
| Profile D: Less interference with daily life | Lewis (17 yrs) | .04 | -.05 | -.05 | **.66** |
|  | Georgia (17 yrs) | -.27 | .19 | .00 | **.65** |
|  | Meghan (21 yrs) | .25 | -.07 | .016 | **.51** |
| Not assigned | Karimah (19 yrs) <sup>a</sup> | -.09 | .55 | .54 | -.09 |
|  | Faizah (18 yrs) <sup>b</sup> | .29 | -.05 | .06 | .04 |
|  | Lien (21 yrs) <sup>b</sup> | .03 | .25 | .11 | -.25 |
| **Variance Explained (%)** |  | 13% | 15% | 14% | 8% |
| **Composite Reliability** |  | 0.96 | 0.97 | 0.97 | 0.92 |

Note: <sup>a</sup> All names are pseudonyms.

**Table 3.** List of Q-set Items with Composite Principal Component Scores (i.e., Ideal-Typical Item Rank)

| # Q-sort item | Item rank |  |  |  |
| --- | --- | --- | --- | --- |
|  | Profile A | Profile B | Profile C | Profile D |
| **Symptoms** |  |  |  |  |
| 1 Being less angry and not losing my temper as much. | -2 | -4 | -1 | 0 |
| 2 Feeling less down and depressed. | **4** | 2 | 2 | **3** |
| 3 Feeling happier and enjoying things more. | **4** | **4** | 1 | **4** |
| 4 Feeling more loved. | **3** | 1 | -2 | -2 |
| 5 Engaging less in behavior that can be harmful. | **3** | -4 | **4** | 0 |
| **Self-management** |  |  |  |  |
| 6 Being more active and engaged in things. | 0 | -2 | 0 | -2 |
| 7 Knowing ways to cope with my emotions. | **3** | **3** | 1 | -4 |
| 8 Having a better understanding of my feelings and thoughts. | 0 | **3** | 0 | -1 |
| 9 Being able to challenge negative thoughts and approach situations differently. | 2 | **4** | 1 | -3 |
| **Functioning** |  |  |  |  |
| 10 Being better able to get things done (e.g. concentrate, be organized). | 1 | 0 | -2 | 0 |
| 11 Being able to do the same things other adolescents do. | -2 | -2 | -3 | **4** |
| 12 Working more effectively in school (e.g. being more motivated and focused). | 0 | -1 | -1 | 0 |
| 13 Attending school more regularly. | -2 | 0 | -3 | -3 |
| 14 Being more sociable and better able to be around other people. | 2 | -1 | -3 | 1 |
| **Personal growth** |  |  |  |  |
| 15 Feeling more confident. | 1 | 0 | 0 | **3** |
| 16 Being better able to stand up for my needs and opinions. | -1 | -3 | 0 | -2 |
| 17 Being more independent and able to take responsibility for my life. | -1 | 2 | 0 | 0 |
| 18 Being able to make sense of things that have happened in the past, or that are still happening. | 1 | 2 | **3** | 1 |
| 19 Having a better sense of who I am and how to be myself around others. | 0 | 1 | -1 | 1 |
| **Relationships** |  |  |  |  |
| 20 Feeling more able to talk about my feelings and thoughts. | -1 | **3** | -1 | 2 |
| 21 Getting on better with my family | 0 | 0 | 2 | -4 |
| 22 Getting on better with my friends or having made new friends. | 1 | -3 | -4 | -1 |
| 23 Getting on better with my peers in school (e.g. not feeling bullied). | -2 | -3 | -4 | -3 |
| **Therapeutic space** |  |  |  |  |
| 24 Having a space where someone listens and cares about me. | -1 | -2 | **3** | -2 |
| 25 Having a space where I can let out my feelings. | -1 | 0 | -1 | 2 |
| 26 Having a space where I can talk about anything without being judged. | 0 | 1 | 1 | -1 |
| 27 Having a space to reflect and think about things differently. | -3 | 0 | 0 | -1 |
| **Well-being** |  |  |  |  |
| 28 Having greater peace of mind (e.g. feeling calmer, more balanced). | 2 | 1 | 2 | -1 |
| 29 Feeling more optimistic and positive about life and the future. | 2 | 2 | **4** | 2 |
| 30 Feeling physically healthier. | 0 | -2 | -2 | 0 |
| 31 Being able to make plans for the future and have goals. | 1 | 0 | 2 | 2 |
| **Parental support and well-being** |  |  |  |  |
| 32 My parents feeling happier and less stressed and worried. | -3 | -1 | 0 | 1 |
| 33 My parents having a better understanding of me and my difficulties. | -3 | 1 | **3** | 0 |
| 34 My parents feeling more able to support me. | -4 | -1 | 1 | 1 |
| 35 My parents feeling less guilty. | -4 | -1 | -2 | **3** |

### Outcome Priority Profile A: Symptoms Reduction and Enhanced Well-Being

Profile A represents six youth and explains 13% of the common variance. A primary desire for this profile was to overcome typical depressive symptoms, such as low mood and affect: “I’d been feeling it for so long it was like something that I just wanted to get rid of. And especially because like ‘feeling less down and depressed,’ for a depressed person that seems like … heaven” (Becca, 17 years).

Young people prioritized *feeling less down and depressed* (item 2, rank +4), *feeling happier and enjoying things more* (item 3, rank +4), *feeling more loved* (item 4, rank +3), *engaging less in behavior that can be harmful* (item 5, rank +3), and *knowing ways to cope with emotions* (item 7, rank +3). Other priority outcomes related to a broader sense of well-being, such as greater *peace of mind* (item 28, rank +2) and *optimism about life and the future* (item 29, rank +2). Youth tended to perceive their depression as a barrier to connecting with others, and ranked *being more sociable* (item 14, rank +2) and *getting on better with friends* (item 22, rank +1) as more important than other outcome profiles. In turn, outcomes related to parental support and parental well-being were deprioritized (items 32– 35, ranks −3 to −4). Some youth preferred to manage their difficulties on their own, while others doubted their parents’ ability to understand and support them.

Of the six adolescents representing this profile, half were female. One was receiving treatment at the time of the research, while five had a history of past treatment. One had been admitted to emergency care in relation with their depression. Youth reported experience with four additional mental health difficulties, on average, including anxiety (*n* = 6), sleep (*n* = 4), self-harm (*n* = 4), and disordered eating (*n* = 4).

### Outcome Priority Profile B: Improved Coping and Self-Management

Profile B represents eight youth and explains 15% of the common variance. Contrary to Profile A, these youth did not believe in an ultimate cure for their depression symptoms, and prioritized gaining the coping skills and agency required to manage their symptoms proactively and independently in the longer term:

> Like when the first wave of sadness hits, normally you don’t have the strategies, you will just […] be snowballing, but […] it’s important to find ways to kind of break that momentum and stop that snowball before it just gets worse. (Jacob, 21 years)
>
> When I first went to CAHMS [child and adolescent mental health services], it was a case of I just wanted to not feel this way anymore. But when I kept going back to CAHMS, then I thought, this isn’t sustainable, I need to be able to function without CAHMS, so the more times I cycled through getting help, the more important sort of resilience or being able to help myself became. (Hannah, 17 years)

Priority outcomes included *feeling happier and enjoying things more* (item 3, rank +4), *being able to challenge negative thoughts and approach situations differently* (item 9, rank +4), *knowing ways to cope with emotions* (item 7, rank +3), *having a better understanding of feelings and thoughts* (item 8, rank +3), and *feeling more able to talk about feeling and thoughts* (item 20, tank +3). Other outcomes considered important included *being able to make sense of past and current experiences* (item 18, rank +2) and *becoming more independent and able to take responsibility for their lives* (item 17, rank +2). The outcomes ranked as least important related to issues that these youth did not primarily struggle with, such as managing anger (item 1, rank −4), reducing harmful behavior (item 5, rank −4), family and peer relationships (items 22 and 23, rank −3), and assertiveness (item 16, rank −3).

Young people in this profile reported the lowest burden from depression and other mental health difficulties, compared with other profiles, with no history of accessing emergency or inpatient care in relation with depression. On average, youth described mental health difficulties in three additional areas, including anxiety (*n* = 5), eating or sleep problems (*n* = 4), and self-harm (*n* = 3). Three out of seven were female.

### Outcome Priority Profile C: Better Understanding Past & Present

Profile C represents eight youth and explains 14% of the common variance. This profile focused on finding safe outlets for emotions, making sense of experiences, and gaining a more positive outlook into the future. Most young people in this profile reported overlapping needs, including learning difficulties (*n* = 4), Autism spectrum disorder (*n* = 3), ADHD (*n* = 2) or a history of trauma (*n* = 2). Youth reported experiencing life differently from others—especially family members—and struggled to make sense of and articulate these experiences. They worried about accessing higher education and employment in the future. These experiences and concerns were reflected in outcome priorities, which included *feeling more optimistic about life and the future* (item 29, rank +4), feeling more understood by their parents (item 33, rank +3) and getting on better with their family (item 22, rank +2), *having a space where somebody would listen and care about them* (item 24, rank +3), *having greater peace of mind* (item 28, rank +3), *and being able to make sense of past and current experiences* (item 18, rank +3),

> I kind of wanna get all my thoughts in order and there’s a lot of stuff that has happened in the past that I wanna deal with before I start dealing with stuff now. (Chelsea, 17 years)
>
> With the Asperger’s, I don’t really understand emotions in general […] I can never tell if I’m sort of truly feeling something or if I’m just thinking I’m feeling that. (Jade, 19 years)

Youth also prioritized *engaging less in behavior that could be harmful* (item 5, rank +4), with self-harm described as an “outlet” for overwhelming emotions. Outcomes around improved peer relationships (items 22 and 23, rank −4), and psychosocial functioning (items 11, 13, 14; rank −3) were assigned low importance. Several youth explained that they did not strive to be “typical” adolescents and felt at ease with a select group of friends or with being on their own.

Seven of the eight youth in this profile were female. Half had completed several courses of treatment, three had visited emergency care in relation with their depression, and one had spent time in inpatient care. On average, they reported five additional mental health difficulties, with anxiety being the most frequent (*n* = 7), followed by self-harm (*n* = 6) and sleep problems (*n* = 5).

### Outcome Priority Profile D: Less Interference with Daily Life

Youth Profile D represented three youth whose struggle with depression and other mental health difficulties had caused substantial impairment, such as having to interrupt school, being unable to go out with friends or to use public transport. As described by Georgia (17 years): “It’s affected everything, like literally everything.” To this group, *feeling happier and enjoying things more* (item 3, rank +4) and *being able to do the same things other adolescents do* (item 11, rank +4) were the two most important outcomes. *Feeling less down and depressed* was also highly ranked (item 2, rank +2). These youth further prioritized recovering confidence and hope (item 15, rank +3; items 29, 31, rank +2) to be able to envisage an identity and future beyond their mental health difficulties:

> Who I am can feel quite dependent on my mood at that moment, and if I’m feeling very low then I’m like […] nothing’s ever gonna be worth it …’ (Meghan, 21 years)

These youth worried about the impact that their mental health difficulties had had on their families. Profile D was the only profile to prioritize *parents feeling less guilty* (item 35, rank +3), and *parents feeling less stressed and worried* (item 32, rank +1). Contrary to other profiles, these youth did not prioritize improvements in coping and self-management (items 6–9, ranks −1 to −4), expressing skepticism that such strategies could be deployed at will, especially when emotions became overwhelming.

Of the three adolescents representing this profile, two were female. Two were receiving ongoing treatment, and two had engaged in several cycles of treatment. Two had been admitted to both emergency care and inpatient care in relation with their depression. These youth reported the highest burden from additional mental health difficulties (in seven areas on average) out of all four profiles. All reported anxiety, self-harm, and problems with eating and sleep. Two each reported a history of psychosis, trauma, and issues with anger and aggression.

### Common Outcome Themes Across Youth Profiles

There was consensus amongst all four outcome priority profiles on the importance of reducing low mood and affect as core depressive symptoms. *Feeling happier and enjoying things more* (item 3) was ranked as *most* important (rank +4) by three out of four profiles, and *feeling less down and depressed* (item 2) was considered important (ranks +2 to +4) by all. Another outcome considered important by all profiles was *feeling more optimistic and positive about life and the future* (item 29, ranks +2 to +4). Most consistently ranked as unimportant was *getting on better with my peers in school* (item 23, ranks −2 to −4), as many young people explained that they did not struggle with peer relationships or bullying.

## Discussion

This Q-study demonstrates plurality in young people’s views about the outcomes that are most important when receiving treatment for depression. We identified four distinct profiles that respectively prioritized outcomes related to symptom reduction and enhanced well-being, improved coping and self-management skills, making sense of past and current experiences, and reduced interference of depression with daily functioning. Despite this heterogeneity, feeling less down and depressed, and enjoying life more were consensus outcomes that were assigned high priority across all four profiles.

The consistent prioritization of improved mood and affect aligns with findings from a qualitative content analysis of change narratives, which found this to be the most salient outcome theme amongst adolescents, parents, and therapists discussing change observed over the course of psychotherapy [16]. In contrast, symptom change was not identified as a principal theme in a qualitative study about notions of “good outcomes” that involved adolescent service users with mixed presenting problems. Symptom relief may be particularly salient for depressed young people, as several studies have found emotional problems to be associated with greater distress, while behavioral disorders tended to be associated with increased functional impairment [53].

Our findings suggest that there are groups of youth that prioritize a range of other outcomes alongside improved mood and affect, in line with outcome themes that have been highlighted in previous research. These include outcomes related to coping and resilience [16, 20, 21, 27, 54, 55], improved family relationships [16, 27, 56–58], feeling heard and understood [16, 59–64], and recovering a sense of hope and optimism [20]. In contrast, youth in this study did not prioritize outcomes around personal growth (e.g., stronger autonomy and identity) which constitute central elements of a client-driven concept of recovery in the adult literature [65], and a key theme in at least one consultation with youth [20]. Similarly, change in peer relationships and friendships did not constitute a priority outcome, in this study although such changes were discussed by a third of young people interviewed following psychotherapy for depression by Krause and colleagues as part of a previous study [16]. This highlights the added degree of differentiation in outcome priorities enabled by the use of Q-methodology.

While Q-methodology is not tailored to the identification of representative socio-economic or clinical profiles, some tentative observations can be made. Young people prioritizing improved coping and self-management skills reported the lowest average burden from depression and other mental health difficulties. They appeared relatively well resourced in terms of individual resilience resources (e.g., confidence, assertiveness) and environmental resources (i.e., support from family and friends), and tended to consider therapy as a temporary additional resource. In contrast, outcomes related to coping and self-management were deprioritized by youth in Profile D who reported the highest burden from mental health difficulties, explaining that their distress would become so overwhelming that deliberate coping was no longer possible or effective. This resonates with previous research suggesting that adults experiencing depression struggle to apply cognitive-behavioral strategies when feeling especially low or overwhelmed [66, 67]. Inversely, Profile D was the only profile to prioritize outcomes related to reduced functional impairment, which tended to be deprioritized by Profile B.

## Implications

Our findings suggest that reducing core depressive symptoms such as low mood and anhedonia are consensus outcomes that are highly valued by most young people. This supports the inclusion of symptom measures in consensus-based core outcome sets that recommend a minimum battery of outcomes to be tracked by all those providing care to youth with depression [25]. At the same time, our findings suggest there is no one-size-fits all outcome prioritization, as youth diverge on the additional outcomes they value the most. If outcome measurement is to be person-centered, services should aim to identify these personal outcomes through conversation and shared decision-making with young people, and track them alongside symptom change [18, 68, 69]. Services may want to consider administering validated standardized scales to track coping skills, family relationships, hopelessness, or global functioning. Personalized outcome measures such as the Goal-Based Outcome Measure [70] or Youth Top Problems [71] provide opportunities for further tailoring [72–74], or may serve as a decision aid to identify outcomes to track through standardized scales [75].

Our findings suggest that young people’s outcome priorities may vary with levels of need for support. In clinical practice, tailored approaches to outcome tracking may be indicated for different needs-based groups [e.g., 76]. Youth with mild or temporary difficulties who mainly require signposting or advice may benefit from the tracking of coping and self-management skills. In turn, tracking hopelessness, functioning, and suicidality may be indicated for youth with higher levels of distress and impairment, overlapping needs, or persistent risk. Research using representative samples is needed to fully understand the linkages between young people’s demographic, cultural, and clinical characteristics, and their outcome priorities.

## Study Limitations

Several limitations must be noted. We employed opportunistic snowball sampling [77] and participants self-selected into the study. Youth volunteers may have differed from the general population by having a particular interest in research. We aimed to mitigate this risk by recruiting from peer-support groups where individuals did not have to proactively contact the research team to arrange participation. Nevertheless, all study participants had successfully navigated their way towards some variation of mental health support and were willing to speak about this experience without fear of stigma. Youth to whom this does not apply may have distinctively different outcome priorities, which may have been missed.

The Q-set was derived using a multi-stage process that drew on the literature and inputs from young people, clinicians, and parents. Nevertheless, the final Q-set represents only a subset of possible outcomes. Additional outcomes that participants suggested could have been included were reduced loneliness and boredom, coming off medication, finding a hobby or an interest, improved general well-being, feeling less guilty to be depressed, improved sleep, being more productive at work, being able to trust others.

Q-methodology is not suited for informing generalizations about the distribution of viewpoints in the wider population [32]. It does, however, allow for generalizations “with respect to the subjectivity at issue”, that is about the existence of the identified profiles in one segment of the population [78]. This study does not claim that the outcome profiles identified are exhaustive or representative of all young people seeking help for depression in the UK, or all professionals supporting them, but provides a basis upon which new and more informed hypotheses can be built [30].

## Conclusions

This study showcased considerable diversity in treatment outcome priorities amongst youth with lived experience of depression, although all tend to give high importance to improvements in mood and affect. Outcomes valued beyond these core outcomes are currently rarely measured in clinical research. This may lead to poor alignment between parameters used to judge treatment efficacy, and the needs and priorities of youth themselves. In clinical practice, ensuring that outcome measurement is acceptable, meaningful, and person-centered requires shared decision-making about the outcomes to track. While core outcome sets can guide the measurement of essential consensus outcomes, an additional element of personalization is likely needed to reflect young people’s individual priorities.

## Data Availability

The data referred to in this manuscript cannot be made publicly available, as study participants did not provide explicit consent for data sharing.

## Acknowledgements

We are grateful to the young people who participated in this study, and to all those who supported the recruitment process. We thank the Anna Freud National Centre for Children and Families in London for lending financial and institutional support to this research, and the Cundill Centre for Child and Youth Depression at the Centre for Addiction and Mental Health in Toronto for funding the first author (KRK) during the finalisation of this manuscript via a post-doctoral fellowship.

## Availability of data and material

N/A

## Code availability

N/A

